# Loss of severe respiratory syncytial virus infection-associated antibody function during the peak of the COVID-19 pandemic mitigation measures

**DOI:** 10.1101/2023.07.30.23292881

**Authors:** Bahaa Abu-Raya, Frederic Reicherz, Christina Michalski, Abdelilah Majdoubi, Liam Golding, Marina Vienta, Madison Granoski, Aleksandra Stojic, David J Marchant, Pascal M. Lavoie

## Abstract

Studies have linked reduced respiratory syncytial virus-specific Fc-mediated phagocytic function and complement deposition to more severe infection. This study shows a loss of these functions during the first year of COVID-19 pandemic. These findings corroborate other data supporting a general waning of RSV antibody functions in absence of viral circulation.

---

Respiratory syncytial virus (RSV) is a major public health concern and a leading cause of morbidity and mortality from lower respiratory tract infection (LRTI) among young children and frail elderly older than 65 years of age [1]. Antibodies protect against RSV disease through antigen-specific, Fab-mediated antibody interactions responsible for virus capture and neutralization [2]. Based on studies in non-human primates challenged with RSV and a human vaccine trial, Fc-mediated antibody functions mediate effector responses against RSV, including phagocytosis of viral particle and complement-deposition, and are important for protection against severe RSV LRTI [3] [4]. Levels of pre-fusion RSV fusion (F) IgG levels and neutralizing titers against RSV A have been shown to decline significantly during the peak of the COVID-19 pandemic measures, during the nearly complete absence of RSV circulation *[5]* [6], suggesting that the duration of the protective antibody immunity to RSV is relatively short-lived. Authors of a modeling study that was informed by epidemiological observations have estimated that RSV immunity against symptomatic re-infection lasts between 6 to 12 months [7]. These observations have clinical implications, indicating that ongoing viral exposure may be required to maintain high antibody levels within populations. Furthermore, while decreased RSV antibody neutralization may explain the increase in symptomatic pediatric RSV cases, it does not explain well the corresponding increase in hospitalizations observed in young children following the relaxing of pandemic measures [8, 9]. This study specifically sought to compare key RSV-specific Fc-mediated functions (i.e., antibody-dependent cellular phagocytosis [ADCP], antibody-dependent complement deposition [ADCD], and antibody-dependent neutrophil phagocytosis [ADNP]) before and after a period of lack of RSV circulation in BC.

## MATERIALS AND METHODS

### Sample description, laboratory analysis and ethics

Analyses were carried out on paired serum samples collected from women 18 to 51 years old during the complete pandemic-related lockdown in BC, Canada, between May to June 2020, and nearly one year later, from February to May 2021, as described (*8)*. ADCP, ADCD and ADNP assays were performed as described (Supplementary Methods, Supplementary Figure 1, Supplementary Figure 2, Supplementary Figure 3). The study was approved by University of British Columbia Children’s and Women’s Hospital Research Ethics Board (H20-01205, H18-01724). All participants provided informed consent for these studies.

### Statistical analysis

Log-transformed data for ADCP, ADCD and ADNP were compared between the 2020 and the 2021 outcomes, by paired t-test. Correlations between fold-changes in ADCP, ADCD and ADNP, and previously published [5] pre-fusion RSV F protein IgG levels and RSV A live virus microneutralization outcomes (expressed antibody titer needed to achieve 95% viral neutralization, or NT95) from the same individuals in 2021 vs 2020 were estimated using Pearson correlation, where statistical significance levels were adjusted for multiple comparisons using the Bonferroni method (p<0.005). To explore functional relationships between RSV antibody outcomes, unsupervised hierarchical clustering was performed. Results for 2020 and 2021 were independently displayed to enable visualization of the relationship between the years and clusters of functions and levels of RSV antibodies. R version 4.1.2 was used for all analyses.

## RESULTS

The median age of the women (n=18) was 37 years (interquartile range [IQR]: 28 – 41 years), with a median of 277 days (IQR: 256–301) between samples collected in 2020 vs. 2021. Both ADCP [geometric means: 228.7 (95% CI: 83.8 – 624.0) vs. 685.5 (95% CI: 338.6-1,388.0); p=0.020] and ADCD [geometric means: 1.26 (95% CI: 0.3 – 5.1) vs. 62.8 (95% CI: 45.3 – 87.2); p<0.001) scores significantly decreased in 2021 compared with 2020 (Figure 1A, 1B). However, no statistically significant difference in ADNP scores were detected between the 2021 and the 2020 samples [geometric means: 2,741 (95% CI: 2,430 – 4-3,091) vs. 2,778 (95% CI: 2,458 – 3,141); p=0.870] (Figure 1C).

**Figure 1:**
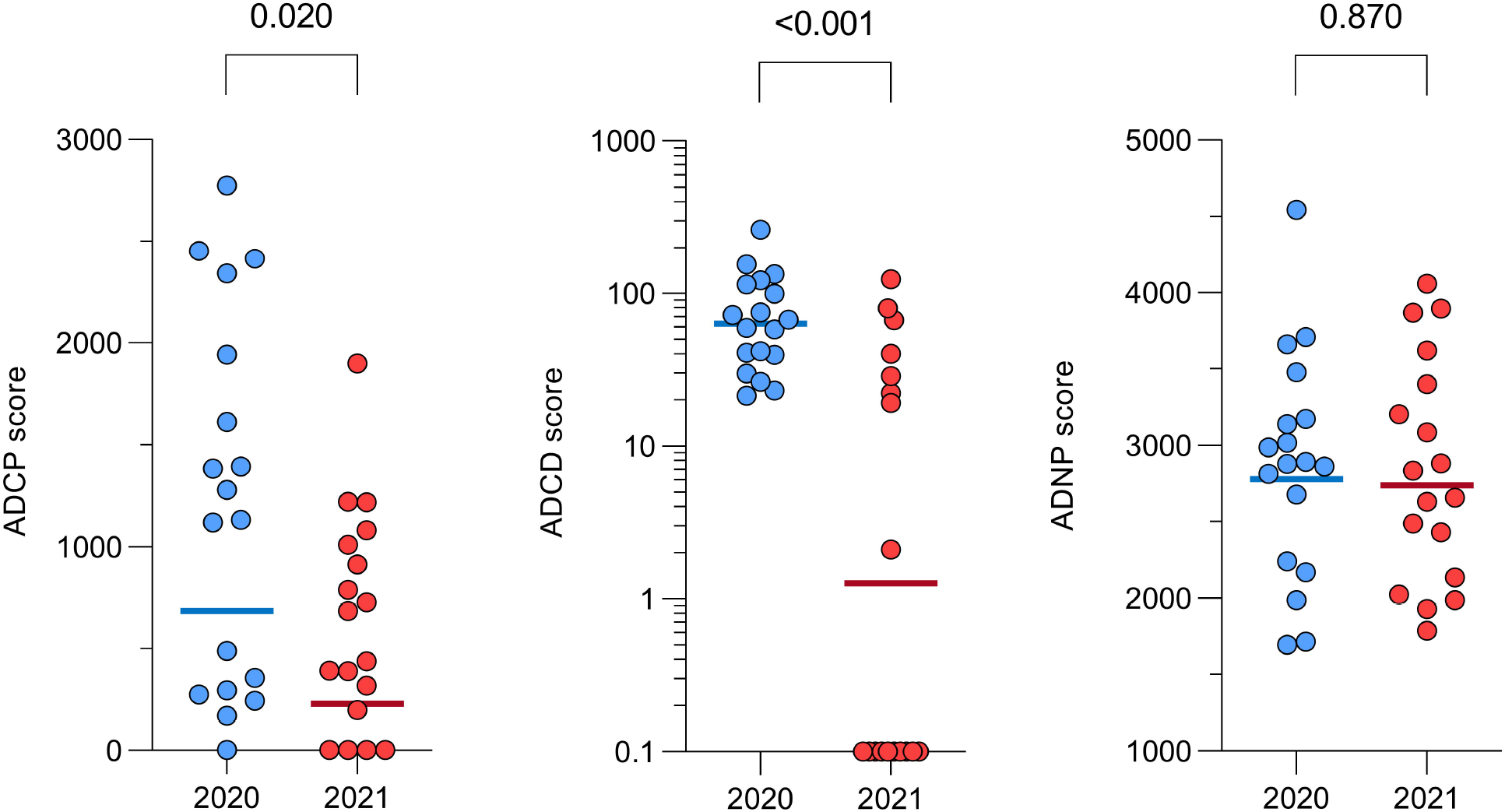
Function of RSV-specific antibodies in paired samples of women aged 18-51 years collected in 2020 (early pandemic) and 2021 (nearly one year after the pandemic) (A-C). **A**) Monocyte antibody-mediated cellular phagocytosis (ADCP); **B**) Antibody-mediated complement deposition (ADCD); **C**) Antibody-mediated neutrophil phagocytosis (ADNP). **Abbreviations:** Pre-F IgG: Prefusion RSV F protein IgG levels; NT95 A: neutralization titer 95 of RSV strain A; ADCP: antibody-mediated cellular phagocytosis; ADCD: Antibody-mediated complement deposition; ADNP: Antibody-mediated neutrophil phagocytosis.

The correlation between fold-changes and levels of functions of RSV antibodies measured in this study (ADCP, ADCD, ADNP) and those we recently published (pre-fusion RSV F protein IgG levels, NT95 RSV A) [5] between 2021 vs 2020 was low to moderate and not statistically indicating that these functions decreased independently of each other and of pre-fusion RSV F protein IgG levels (Supplementary Figure 4, Supplementary Figure 5). The samples collected in 2020 and 2021 showed a separate unique cluster that displayed lower ADCP and a unique subset of subjects with lower ADCD using unsupervised hierarchical clustering (Supplementary Figure 6).

## DISCUSSION

This study adds to recent evidence, supporting a broad decline in RSV antibody functions beyond antibody-mediated viral neutralization. The waning of Fc-mediated antibody functions essential for cellular responses and associated with RSV LRTI may herald more severe RSV outcomes, and not just an increase in symptomatic RSV cases, as suggested in epidemiological studies [8, 9]. Indeed, reduced RSV antibody outcomes in women of childbearing age could impair the degree of RSV antibody immunity transferred to infants born during this period. The waning of Fc-mediated antibody functions may also result in increased vulnerability in older children who were unexposed to RSV repeatedly in a similar period. Moreover, the deficit in RSV antibody outcomes observed across multiple immunological functional domains strengthen the evidence for an increase pool of immunologically susceptible individuals during the strict period of application of pandemic measures, especially in immunologically naïve infant who lack immunological protection from acquired RSV T cell memory at this young age.

Examining RSV antibody functions in paired serum samples allowed to minimize the individual variability expected for these outcomes in humans [10]. ADNP and ADCD correlated with a reduction in viral load in bronchoalveolar lavage and thus protection from LRTI disease in non-human primates challenged with RSV [3]. ADCP, ADNP and ADCD correlated with protection from symptomatic RSV infections in humans vaccinated with the adenovirus-based Ad26.RSV.preF vector [4]. Based on these studies, phagocytosis of RSV immune complexes effectively activates target cells, appear necessary for a broad range of effector immune responses [11], indicating that RSV protection cannot simply be assessed solely by measuring RSV antibody levels [10]. Consistent with this, while we showed that antibody-mediated phagocytosis declined using monocytes as the target cells (in the ADCP assay), this was not shown using neutrophils as the target cells (in the ADNP assay). This finding could be due to differential expression of receptors implicated in the function on target cells and their affinity to RSV F IgG, and the collaboration between Fab-mediated and Fc-mediated functions [12, 13]. Immune system cells (e.g., monocytes, neutrophils) express Fc receptors with both activation and inhibition functions and the balance of these opposed signal pathways is important in regulating each effector response. THP-1 monocytes express FcγRII (low affinity [FcγRIIA activating and FcγRIIB inhibitory]) at higher levels than FcγRI (high affinity, activating) and FcγRIII (low affinity, activating) [14]. FcγRIIA is the isoform implicated in phagocytosis of antigen-antibody complexes. Each neutrophil expresses 30,000 to 60,000 copies of FcγRIIA (low affinity), a level that is less abundant than FcγRIIIB [12, 13]. It is thus possible that high expression of FcγRII makes THP-1 cells more sensitive in capturing changes in phagocytosis activity of IgG. These findings highlight the importance of examining multiple immune outcomes to obtain a more complete picture of RSV antibody functions in humans.

Our study is limited by its small sample size and performed in a single region and the lack of parallel clinical outcome measures. Notably, BC, experienced significant increase in RSV cases with increased hospitalizations in older children after the relaxing of pandemic measures, providing direct population correlates supporting the clinical relevance of the current findings (*9*). Overall, our study sheds light on the longevity of, and provides further evidence for a waning of RSV antibody outcomes associated with more severe RSV infection. Further studies are needed to replicate these findings in other health jurisdictions.

## Data Availability

All data produced in the present study are available upon reasonable request to the authors

## Acknowledgments

The authors wish to acknowledge BC Children’s Hospital Research Institute Core Technologies and Services, Vancouver, Canada for providing the support to perform the flow cytometry experiments. The authors would like to thank Dr. Jeffrey Bone, Biostatistical Lead at BC Children’s Hospital Research Institute for providing input on statistical analysis.

## Author contributions

Conceptualization: BA, FR, PML

Methodology: Neutralization of RSV A (FR); ADCP, ADNP and ADCD (BA with input from CM, PML)

Data analysis and visualization: BA, FR

Data interpretation: BA, CM, FR, MV, PML, AM, LG, MG, AS, DM

Writing – original draft: BA

Writing – review & editing and approval of submitted paper: BA, CM, FR, MV, PML, AM, LG, MG, AS, DM

