## SupplementaryMaterial for "Loss of severe respiratory syncytial virus infection-associated antibody function during the peak of the COVID-19 pandemic mitigation measures"

**Supplementary Material**

**Supplementary Methods**

**Supplementary Figure 1:** Gating strategy for antibody-dependent cellular phagocytosis on singlet live cells.

**Supplementary Figure 2:** Gating strategy for antibody-dependent neutrophil phagocytosis on CD66b+ singlet live cells.

**Supplementary Figure 3:** Gating strategy for antibody-dependent complement deposition on singlet red beads.

**Supplementary Figure 4:** Scatter correlation plots of fold-changes in functions of RSV antibodies of women aged 18-51 years.

**Supplementary Figure 5:** Scatter correlation plots of levels of functions of RSV antibodies of women aged 18-51 years.

**Supplementary Figure 6:** Heat-map analysis based on hierarchical unsupervised clustering of functions of RSV antibodies.

**Supplementary Methods**

Blood sample collection and processing

Blood was collected in gold-top serum separator tubes with polymer gel (BD Biosciences) and left at least 30 minutes at room temperature. Collected blood was centrifuged at 1400 x g for 10 minutes and serum was collected and frozen at ‑80°C within 4 hours of collection.

**Phagocytosis assays:** Antibody-dependent cellular phagocytosis (ADCP) and antibody-dependent neutrophil phagocytosis (ADNP) assays were performed as previously published (*1-3*).

**Antigen-beads coupling:** RSV Fusion protein (Sino Biological, 11049-V08B-00) was biotinylated on lysine residues using a NHS-LC-biotin reagent (Thermofisher Cat#21336) according to the manufacturer's instructions. Biotinylated antigen was incubated with 1 µm fluorescent beads (Thermo Scientific, F8776). Beads were then washed in 0.1% PBS-BSA twice. Washed beads were resuspended with 0.1% PBS-BSA, combined with biotinylated antigen and incubated 2 hours in the dark at 37 °C. Antigen-coated beads were then washed with 0.1% PBS-BSA and resuspended in 0.1% PBS-BSA.

**Antigen-beads incubation with antibodies:** 10 µL of antigen-coated beads were incubated with an equal volume of either heat-inactivated (56 °C for 30 minutes) serum or Palivizumab (positive control) diluted in 0.1% PBS-BSA or PBS (negative control) in a 96-well U-bottom culture plate at 37 °C, 5% CO2 for 2 hours. The immune complexes were spun down and washed in PBS and incubated with THP-1 cells (for ADCP) or neutrophils (for ADNP).

**Monocyte ADCP:** Immune complexes were incubated with 25,000 THP-1 cells (grown in R10 [RPMI plus 10 % fetal bovine serum, sodium pyruvate and penicillin/streptomycin]) per well at a concentration of 1.25 x10^5^ cells/mL in R10 overnight at 37 C, 5% CO_2_. After the incubation, the cells were stained for viability (Thermo Scientific, 65-0865-14) and fixed in 4% paraformaldehyde (Fisher Scientific, 50-980-491). Data were collected on a Fortessa flow cytometer performed at BC Children’s Hospital Research Institute (BCCHR) Core Technologies and Services (Vancouver, Canada) and analyzed using Flowjo (version 10.8.1). Phagocytosis score was calculated as the percentage of bead positive cells, multiplied by geometric mean fluorescence intensity of bead positive cells normalized for the negative control (**Supplementary Figure 1**).

**ADNP:** White blood cells (WBCs) were isolated from freshly drawn peripheral blood by lysing erythrocytes in ACK lysing buffer (Thermo Scientific, A1049201) for 5 minutes. WBCs were pelleted by centrifugation and washed with PBS twice. Cells were finally resuspended at 2.5x10^5^ cells/mL in R10 and 50,000 cells per well were incubated with immune complexes (as described above) for 1 hour at 37 °C, 5% CO2. Neutrophils were stained with anti-CD66b- Pacific blue (Thermo Scientific, 17-0666-42) and viability dye (Thermo Scientific, 65-0865-14) and fixed with 2 % paraformaldehyde (Fisher Scientific, 50-980-491). Phagocytosis scores were calculated as above in ADCP assay based on CD66b+ viable cells (**Supplementary Figure 2**).

**Antibody-dependent complement deposition (ADCD):** High throughput ADCD assay was performed as previously published (4). In brief, biotinylated RSV antigen was incubated with 1μm red fluorescent neutravidin beads (Thermo Scientific, F8775) and prepared as described for phagocytosis assays. 10 µL of antigen-coated beads were incubated with an equal volume of heat-inactivated (56 °C for 30 minutes) serum diluted in 0.1% PBS-BSA or PBS (negative control) in a 96-well U-bottom culture plate at 37 °C for 2 hours. The immune complexes were spun down and washed in PBS and incubated with guinea pig complement (Cedarlane, CL4051) reconstituted in R10 (RPMI with 10% FBS) at 37 ° C for 15 minutes. After the incubation, the bead-based immune complexes with complement were spun down and washed twice with 15 mM EDTA in PBS (Invitrogen, AM9260G). Fluorescein-conjugated goat anti-guinea pig complement C3 (MP Biomedicals, cat|# 0855385) was added to the complex and incubated at room temperature for 15 min. Data were collected on a BD LSR II flow cytometer and analyzed using Flowjo (version 10.8.1), with the median fluorescence intensity of all bead positive events in the FITC channel as the readout normalized for the negative control (Supplementary Figure 3).

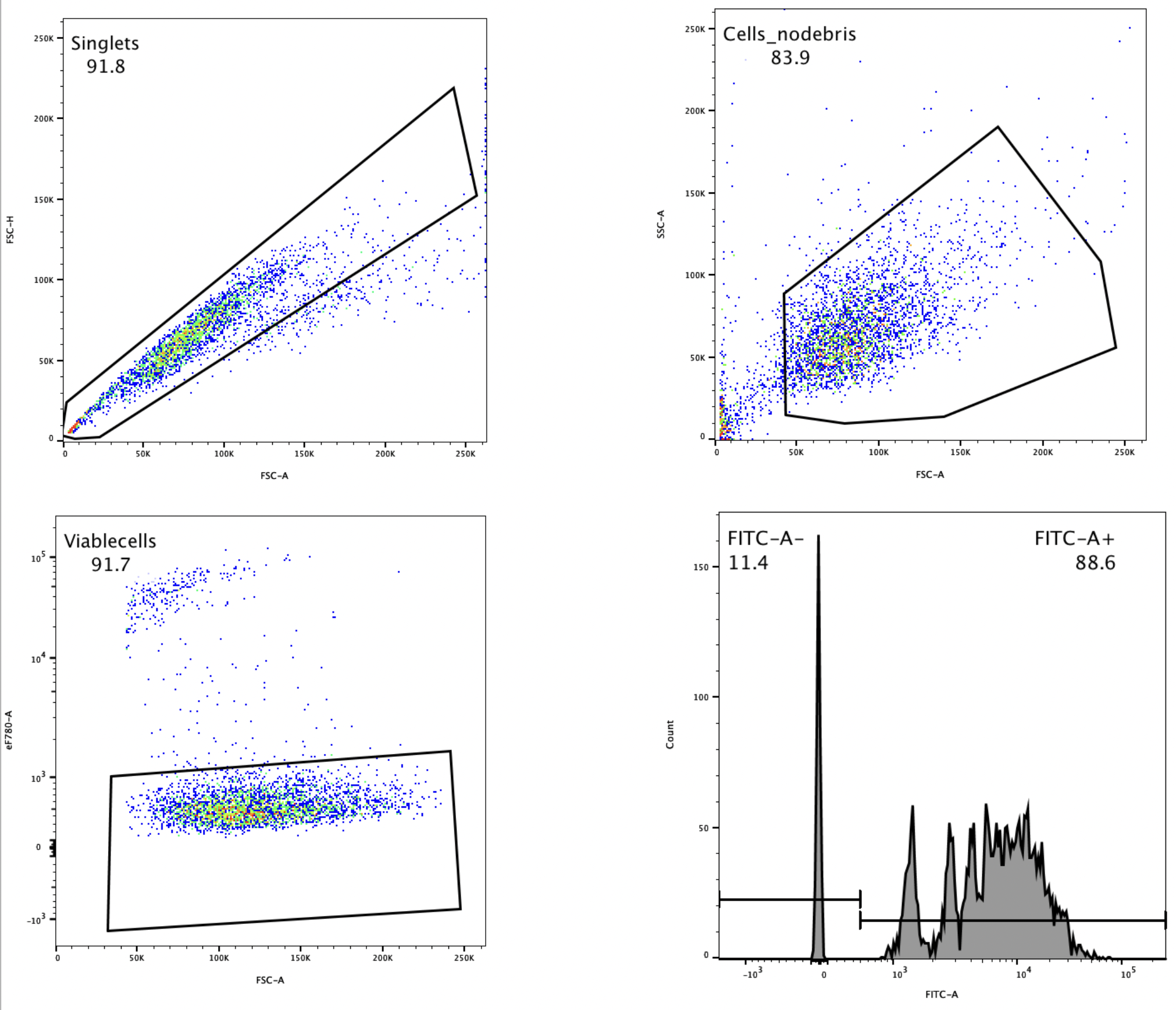

**Supplementary Figure 1:** Gating strategy for antibody-dependent cellular phagocytosis on singlet live cells.

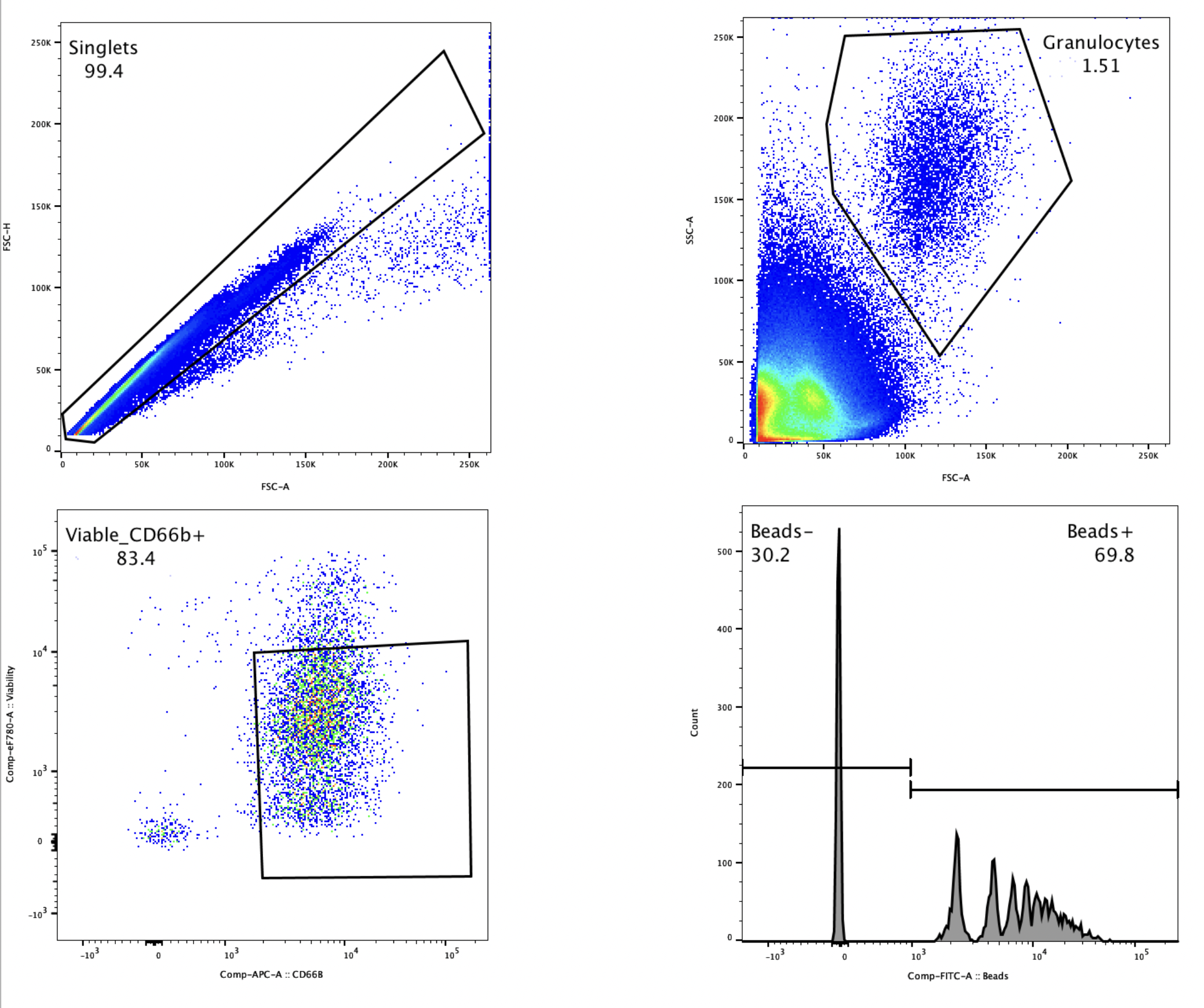

**Supplementary Figure 2:** Gating strategy for antibody-dependent neutrophil phagocytosis on CD66b+ singlet live cells.

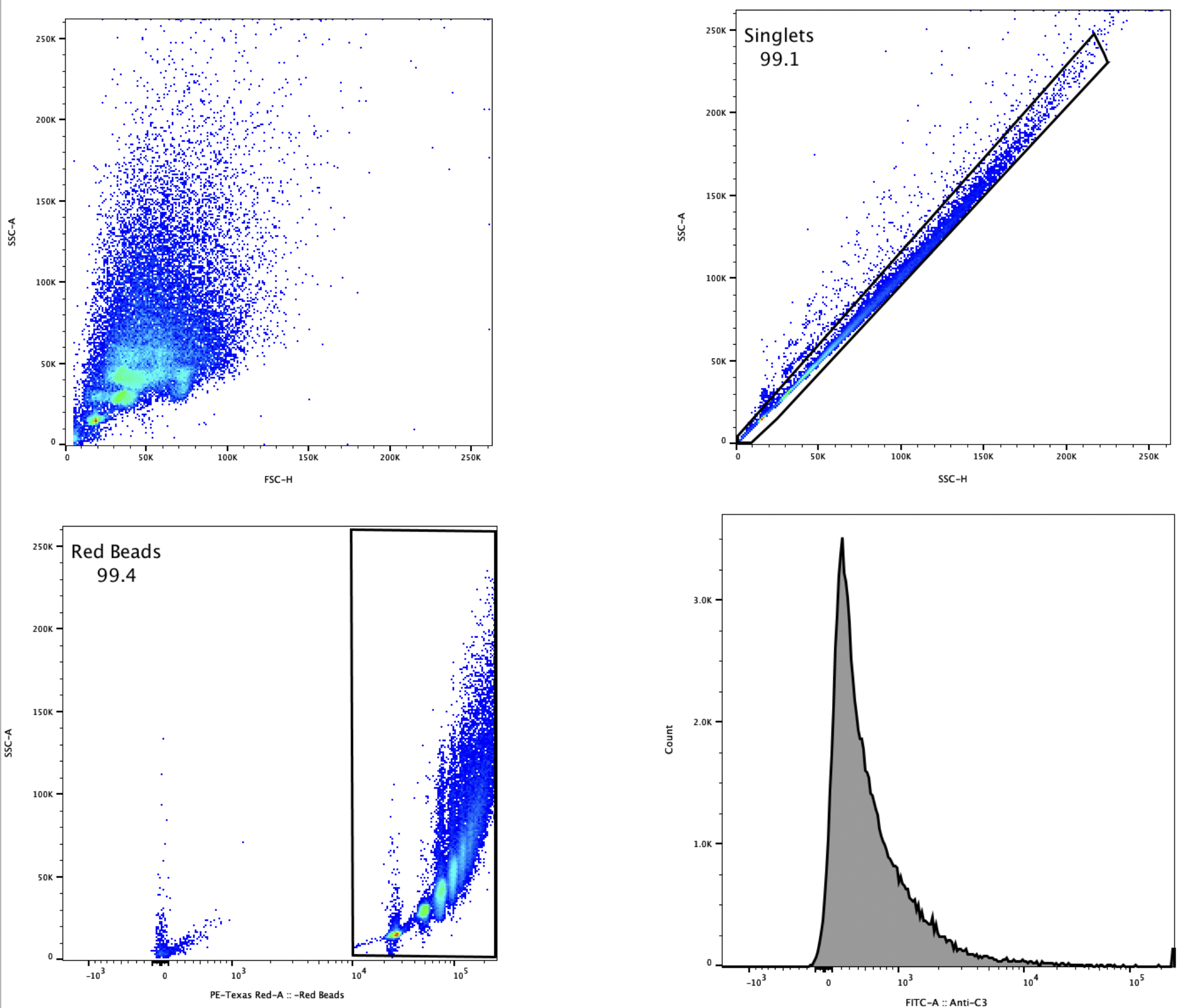

**Supplementary Figure 3:** Gating strategy for antibody-dependent complement deposition on singlet red beads.

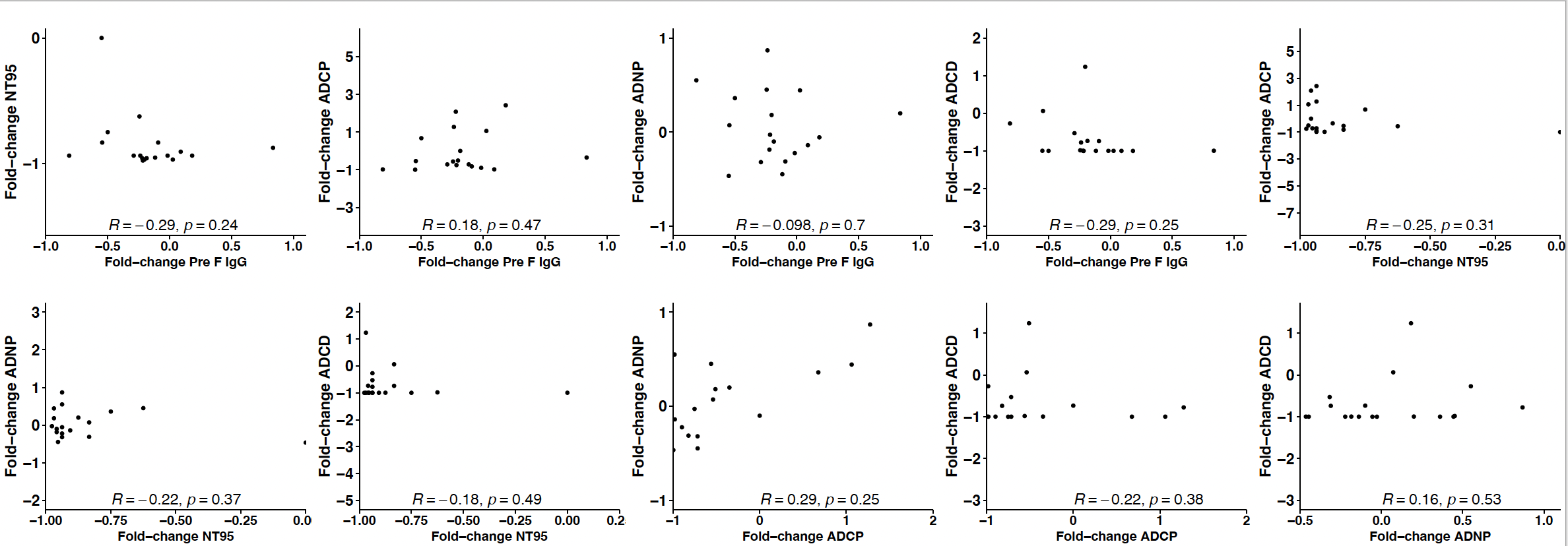

**Supplementary Figure 4:** Scatter plots of correlation of fold-changes in functions of RSV antibodies of women aged 18-51 years. Scatter plots of fold changes of pair of functions of RSV antibodies in all subjects (n=18) based on Pearson’s correlation coefficient test. P-value of 0.05 was assigned and statistical significance of each correlation was set using Bonferroni correction for multiple testing (p<0.005). Abbreviations: Pre F IgG: Prefusion RSV F protein IgG levels; NT95: neutralization titre 95 of RSV strain A; ADCP: antibody-mediated cellular phagocytosis; ADCD: Antibody-mediated complement deposition; ADNP: Antibody-mediated neutrophil phagocytosis.

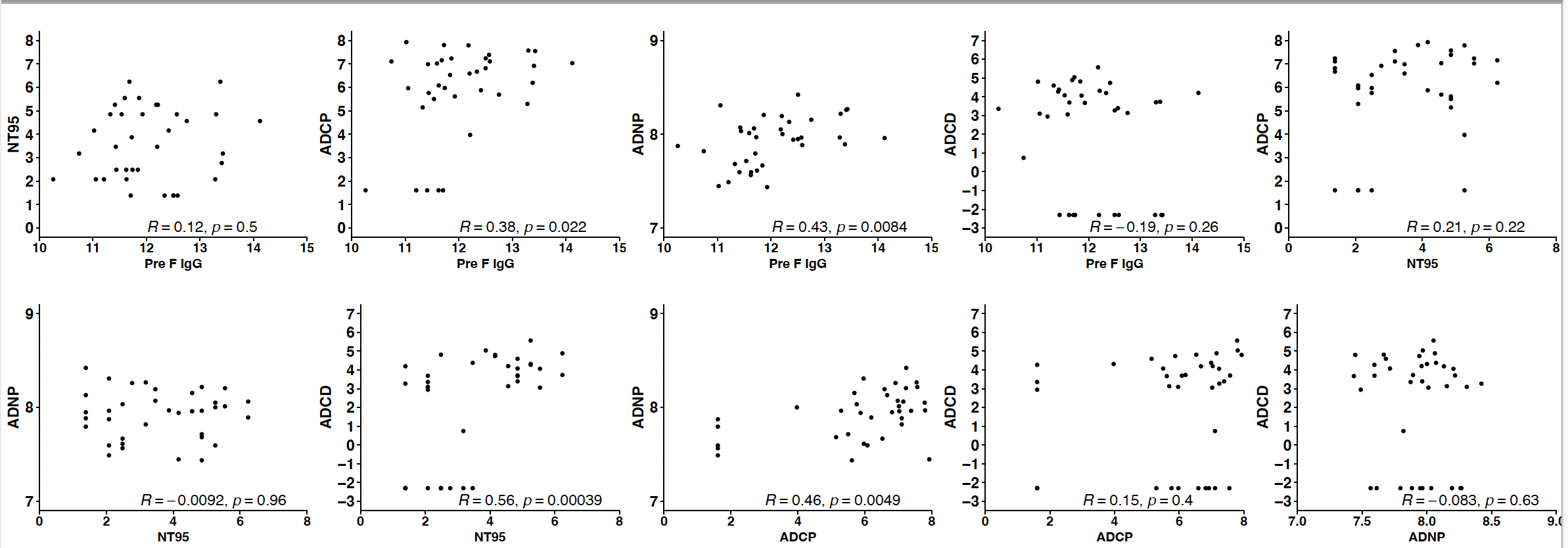

**Supplementary Figure 5:** Scatter plots of correlation of levels of functions of RSV antibodies of women aged 18-51 years. Scatter plots of logged levels of pair of functions of RSV antibodies in all subjects (n=18) based on Pearson’s correlation coefficient test. P-value of 0.05 was assigned and statistical significance of each correlation was set using Bonferroni correction for multiple testing (p<0.005). Abbreviations: Pre F IgG: Prefusion RSV F protein IgG levels; NT95: neutralization titre 95 of RSV strain A; ADCP: antibody-mediated cellular phagocytosis; ADCD: Antibody-mediated complement deposition; ADNP: Antibody-mediated neutrophil phagocytosis.

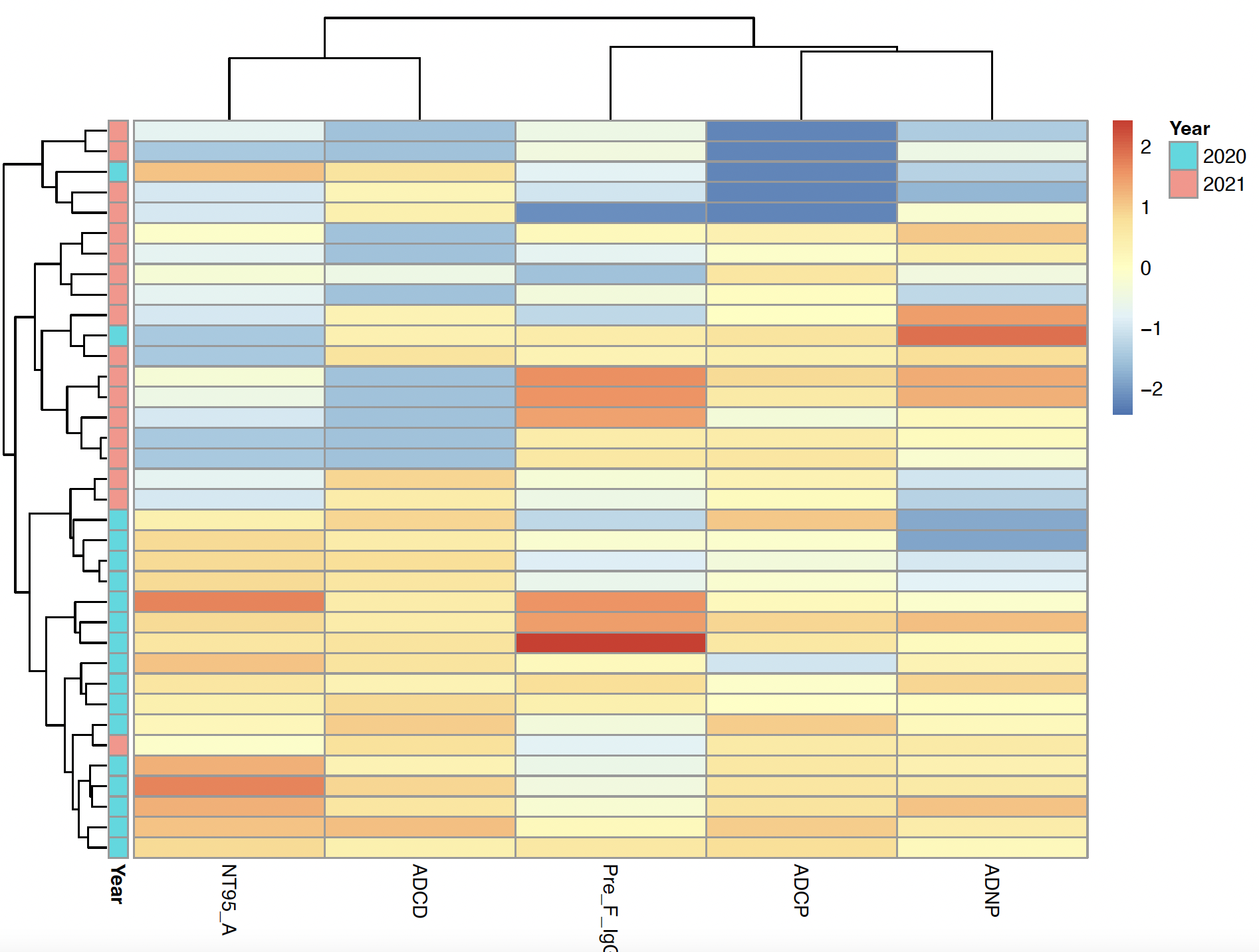
 Supplementary Figure 6: Heat-map analysis based on hierarchical unsupervised clustering of functions of RSV antibodies. The scaled levels (Z-scores) were color-coded as indicated by the scale on the right, in which levels range from blue to red indicating high (red) and low (blue) levels. The years (2020 and 2021) are displayed by a column. Abbreviations: Pre-F IgG: Prefusion RSV F protein IgG levels; NT95 A: neutralization titer 95 of RSV strain A; ADCP: antibody-mediated cellular phagocytosis; ADCD: Antibody-mediated complement deposition; ADNP: Antibody-mediated neutrophil phagocytosis.
